# Point-of-Care CRISPR-Cas-Assisted SARS-CoV-2 Detection in an Automated and Mobile Droplet Magnetofluidic Device

**DOI:** 10.1101/2021.01.27.21250564

**Authors:** Fan-En Chen, Pei-Wei Lee, Joon Soo Park, Alexander Y. Trick, Liben Chen, Kushagra Shah, Kuangwen Hsieh, Tza-Huei Wang

## Abstract

In the fight against COVID-19, there remains unmet needs in developing point-of-care (POC) diagnostic testing tools that can rapidly and sensitively detect the causative SARS-CoV-2 virus to control disease transmission and improve patient management. Although recent CRISPR-Cas-assisted SARS-CoV-2 detection assays (such as DETECTR and SHERLOCK) are viewed as transformative solutions for POC diagnostic testing, their lack of simple sample processing and full integration within an automated and portable device hamper their potential for POC use. We report herein POC-CRISPR – a new single-step CRISPR-Cas-assisted assay that is coupled to droplet magnetofluidics (DM) – that leverages simple magnetic concentration and transport of nucleic acid-binding magnetic beads to accomplish sample preparation and assay automation. By further adapting the assay into a fully integrated thermoplastic cartridge within a palm-sized mobile device, POC-CRISPR was able to detect 1 genome equivalent (GE)/µL SARS-CoV-2 RNA from a sample volume of 100 µL in 30 min. Moreover, when evaluated with unprocessed clinical nasopharyngeal (NP) swab eluates, POC-CRISPR identified SARS-CoV-2 positive samples in as short as 20 min and achieved full concordance with standard RT-qPCR.

## Introduction

Since its initial report in December 2019, COVID-19 has rapidly spread across 220 countries and territories across the globe^[1]^. As of December 15, 2020, there are more than 71 million confirmed cases and more than 1.6 million deaths worldwide.^[1]^ In the fight against this once-in-a-lifetime pandemic, diagnostic testing that can rapidly and sensitively detect the causative virus SARS-CoV-2 at the point of care (POC) is identified by the WHO as a top priority^[2]^ due to the potential for controlling disease transmission and improving patient management. Responding with unprecedented urgency, the global scientific community has made remarkable progress in developing promising tools toward POC diagnostic testing of SARS-CoV-2. This is especially true in the commercial sector, where Abbott’s ID NOW COVID-19 test, which can report results in 5−15min, and Cepheid’s Xpert Xpress SARS-CoV-2, which can report results in 45 min, represent the leading commercial instruments in POC diagnostic testing. Despite these promising developments and several evaluation studies,^[3–8]^ however, there has yet to see wide adoption of these devices or POC diagnostic testing of SARS-CoV-2 in general. Such a gap signals that there remains a significant room for new solutions.

CRISPR-Cas-assisted diagnostic assays (*e*.*g*., SHERLOCK^[9]^ and DETECTR^[10]^) have in recent years captured significant attention.^[11–14]^ Aided by the elegant detection mechanism, these assays have demonstrated fast detection while obviating instrument-intensive thermocycling. As a result, CRISPR-Cas-assisted SARS-CoV-2 detection assays^[15–20]^ are viewed as transformative solutions for POC diagnostic testing. Developing a POC diagnostic test from this nascent method, however, still must address numerous technical requirements. For example, to ensure ease of use, portability, and resource/infrastructure independence, CRISPR-Cas-assisted assays still must reduce assay steps, obviate manual operation in sample processing, and integrate with automated and portable devices. To this end, despite their seminal status, both DETECTR-based^[15]^ and SHERLOCK-based SARS-CoV-2 detection^[16]^ require multiple manual assay steps and input SARS-CoV-2 RNA that is already extracted and purified by benchtop instruments. More recent assays^[21–26]^ have made significant advances but have yet to fully meet these requirements. For example, AIOD-CRISPR^[21]^ (a novel one-step CRISPR-Cas12a-assisted transcription recombinase polymerase amplification^[27]^ (RT-RPA) assay) and the amplification-free CRISPR-Cas13a assay^[25]^ can be detected via a miniaturized detector or a mobile phone, but both still require benchtop-extracted SARS-CoV-2 RNA as the input and separate heating modules for incubation. STOPCovid.v2,^[22]^ a new SHERLOCK assay that couples one-step CRISPR-Cas12b-assisted reverse transcription loop-mediated isothermal amplification^[28]^ (RT-LAMP) to benchtop magnetic-based RNA extraction, represents perhaps the most mature CRISPR-Cas-assisted assay to date, but still relies on manual operation to process samples. Thus, to date, there had yet to be a CRISPR-Cas-assisted SARS-CoV-2 detection assay that is tenable for POC use.

In response, we have developed POC-CRISPR – the first one-step CRISPR-Cas-assisted assay that is fully integrated with sample preparation and implemented in a POC-amenable device. Sample preparation in POC-CRISPR is achieved through droplet magnetofluidics (DM),^[29–35]^ which uses magnetic-based capture and transport of nucleic acid-binding magnetic beads to automate complex sample preparation procedures. DM-enabled sample preparation offers an effective means for concentrating and purifying SARS-CoV-2 RNA from a large volume of swab eluate, as well as for transporting the RNA to downstream amplification and detection. Moreover, we have established a DM-compatible, one-step, fluorescence-based CRISPR-Cas12a-assisted RT-RPA assay for SARS-CoV-2 detection. To facilitate POC use, we have implemented POC-

CRISPR within a plastic assay cartridge that is comparable to a USB stick in size and a lateral flow strip in material cost, and we have developed an automated, palm-sized, mobile device for performing the assay. POC-CRISPR could detect 1 genome equivalent (GE)/µL SARS-CoV-2 RNA from a sample volume of 100 µL in 30 min. Moreover, when evaluated with 8 unprocessed NP swab eluates, POC-CRISPR could identify the 4 SARS-CoV-2 positive samples in 20 – 40 min and achieved full concordance with standard benchtop RT-qPCR.

## Results

### Overview of POC-CRISPR

POC-CRISPR detects SARS-CoV-2 virus directly from unprocessed clinical NP swab eluates in a sample-to-result workflow powered by the integrated mobile DM device (**Figure 1A**). POC-CRISPR begins by injecting the swab eluate and a magnetic bead buffer into an assay cartridge and mounting the cartridge in the mobile DM device. This process takes < 1 min hands-on time and represents the only manual process in POC-CRISPR. Within the mobile DM device, a motorized magnetic arm, a miniature heating module, and a compact fluorescence detector are programmed by a microcontroller to execute sample preparation, reaction incubation, and fluorescence measurements in full automation. Briefly, the motorized magnetic arm first concentrates the magnetic beads that capture SARS-CoV-2 RNA from the swab eluate and then sequentially transports the concentrated beads and bound RNA to the wash buffer well and the CRISPR-Cas12a-assisted RT-RPA reaction mixture well. Upon the arrival of RNA, the heating module heats the reaction mixture well to initiate the reaction while the fluorescence detector periodically measures the fluorescence emitted from the reaction. The resulting real-time fluorescence curves signal the presence or absence of SARS-CoV-2 virus in the NP swab eluate.

**Figure 1.**
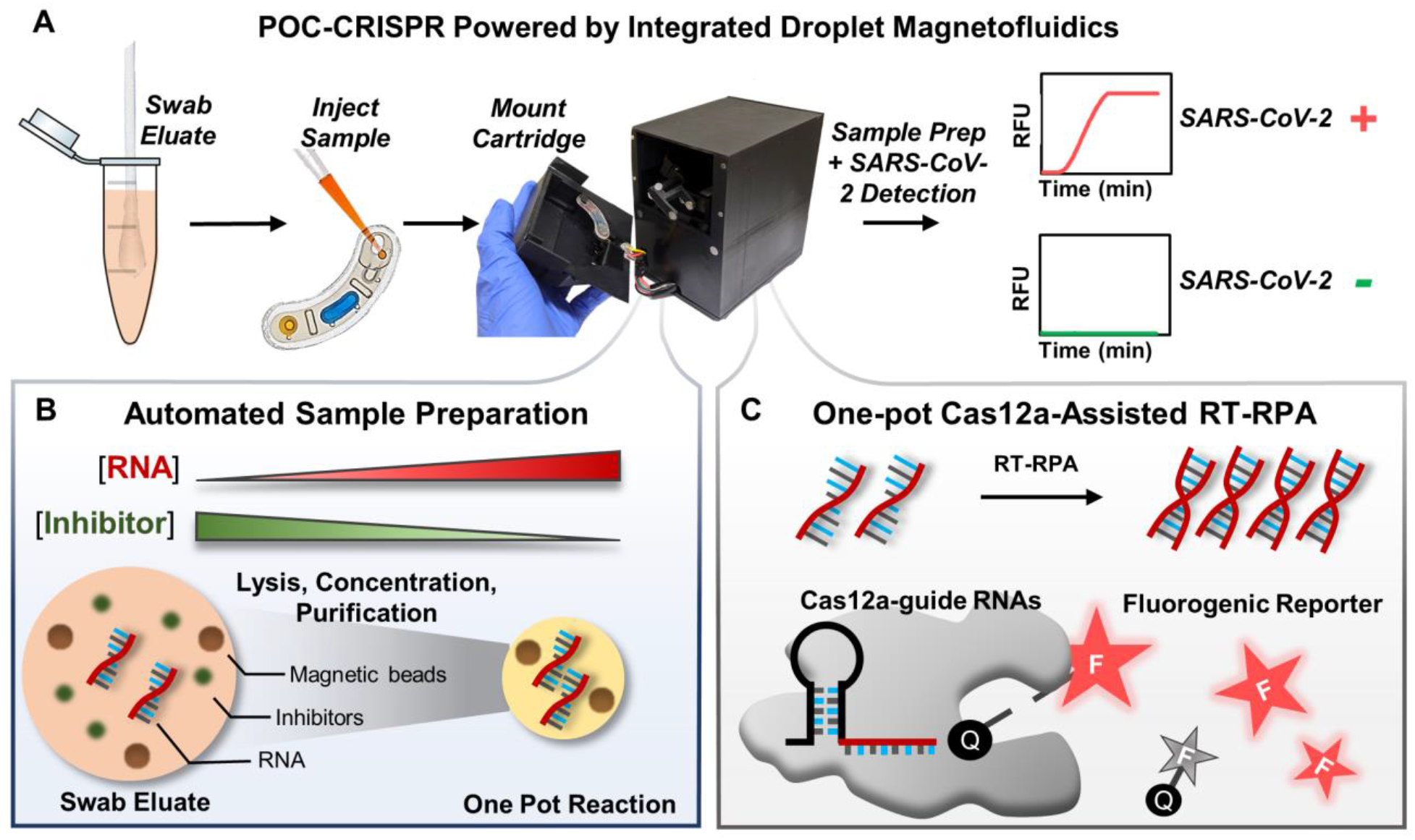
Detection of SARS-CoV-2 from nasopharyngeal swab eluates via POC-CRISPR empowered by integrated droplet magnetofluidics. (A) POC-CRISPR – the first CRISPR-Cas-assisted assay integrated with sample preparation and implemented in a POC-amenable device – can detect SARS-CoV-2 directly from unprocessed nasopharyngeal (NP) swab eluates in a sample-to-result workflow. Upon injecting the NP swab eluate into an assay cartridge and mounting the cartridge into a palm-sized, mobile digital magnetofluidic (DM) device, the device performs sample preparation, reaction incubation, and fluorescence measurements in full automation. The resulting real-time fluorescence curves signal the presence or absence of SARS-CoV-2 virus in the NP swab eluate. (B) Within the device, sample preparation is powered by DM, which leverages magnetic-based capture and transport of nucleic acid-binding magnetic beads to concentrate and purify SARS-CoV-2 RNA from a large volume of NP swab eluate and to transport the RNA into a CRISPR-Cas-assisted reaction mixture. (C) The reaction mixture incorporates reverse transcription recombinase polymerase amplification (RT-RPA) and Cas12a-based collateral cleavage of single-stranded DNA fluorogenic reporter in one pot. Within the reaction, SARS-CoV-2 RNA is *in vitro* transcribed and amplified via RT-RPA into DNA amplicons, which activate Cas12a-guide RNA complexes to cleave single-stranded DNA fluorogenic reporters, thereby producing fluorescent signals for detection.

DM-enabled sample preparation and CRISPR-Cas12a-assisted RT-RPA represent the two crucial assay components in POC-CRISPR. In this work, DM-enabled sample preparation from clinical NP swab eluates is facilitated by pH-mediated electrostatic attraction between RNA and functionalized magnetic beads. Briefly, the swab eluate is first exposed to a low-pH binding buffer supplemented with a detergent, in which the viral particles are lysed and negatively-charged RNA can electrostatically bind to magnetic beads with positive surface charges. As the magnetic beads can be concentrated, they offer an effective strategy for concentrating RNA from large volumes of swab eluates and boosting the assay sensitivity. When the magnetic beads and bound RNA are magnetically transported out of the swab eluate, potential inhibitors to CRISPR-Cas12a-assisted RT-RPA are left behind. Moreover, as the magnetic beads and bound RNA are magnetically transported into a neutral-pH wash buffer, RNA remains bound to the magnetic beads while additional potential inhibitors adsorbed to the magnetic beads are released into the wash buffer, thereby achieving further purification. As a result of DM-enabled sample preparation, concentrated and purified RNA can be delivered to the CRISPR-Cas12a-assisted RT-RPA reaction mixture (**Figure 1B**). In this work, SARS-CoV-2 RNA is detected via a fluorescence-based, one-step CRISPR-Cas12a-assisted RT-RPA reaction. Within this reaction, SARS-CoV-2 RNA is *in vitro* transcribed and amplified via RT-RPA into DNA amplicons, which activate Cas12a-guide RNA complexes to cleave single-stranded DNA (ssDNA) fluorogenic reporters, thereby producing fluorescent signals for detection (**Figure 1C**).

### Development of benchtop DM-compatible CRISPR-Cas12a-assisted RT-RPA

As a prerequisite for realizing POC-CRISPR, we first developed a DM-compatible CRISPR-Cas12a-assisted RT-RPA on benchtop. That is, the CRISPR-Cas12a-assisted RT-RPA can successfully amplify and detect SARS-CoV-2 RNA that is captured and concentrated by magnetic beads. As the testbed, we employed standardized synthetic SARS-CoV-2 RNA from NIAID BEI Resources (NR-52358) as the target. Consistent with our previous DM devices,^[33–34]^ we employed ChargeSwitch magnetic beads and the associated binding buffer as our magnetic bead buffer. For SARS-CoV-2 amplification and detection, we used a fluorescence-based CRISPR-Cas12a-assisted RT-RPA adopted and modified from AIOD-CRISPR^[21]^ and our recently developed deCOViD^[23]^ that used a more sensitive, Alexa647 fluorophore-based ssDNA fluorogenic reporter^[36]^ (Supplementary Information Table S1). For developing this new assay, we mixed SARS-CoV-2 RNA with the magnetic bead buffer and used a benchtop magnetic rack to capture and wash the magnetic beads and bound RNA. We next added the CRISPR-Cas12a-assisted RT-RPA reaction mixture and used a benchtop real-time PCR instrument to incubate the reaction and measure the fluorescence. We finally analyzed the real-time fluorescence amplification curves from the reactions to assess the reaction conditions. We note that, aside from differences in assay components and conditions, our benchtop DM-compatible CRISPR-Cas-assisted SARS-CoV-2 detection assay shares a comparable workflow as STOPCovid.v2^[22]^.

We investigated a range of parameters in our benchtop DM-compatible CRISPR-Cas12a-assisted RT-RPA assay to improve its performance in SARS-CoV-2 detection. In these investigations, we used 100 µL of 100 GE/µL RNA as the sample – up to 100-fold greater in sample volume than typical CRISPR-Cas-assisted assays in which ~1 – 5 µL samples are spiked directly into the reaction mixtures – because magnetic beads could capture and concentrate RNA from such a large sample volume. We established our first version of DM-compatible CRISPR-Cas12a-assisted RT-RPA by incorporating WarmStart RTx reverse transcriptase in the assay, which could be achieved without requiring a designated step for removing the magnetic beads from the reaction mixture (Supplementary Information Figure S1). We then determined that, while multiple reverse transcriptases supported DM-compatible CRISPR-Cas12a-assisted RT-RPA, WarmStart RTx reverse transcriptase allowed the fastest and most efficient detection among the reverse transcriptases we tested (Supplementary Information Figure S2). As WarmStart RTx reverse transcriptase reacts optimally at 55 °C, we next tuned the reaction temperature and found that our RT-RPA was the most efficient at 43 °C and 45 °C (Supplementary Information Figure S3). Finally, we adjusted the concentrations of WarmStart RTx reverse transcriptase, Cas12a, Cas12a-guide RNAs, and ssDNA fluorogenic reporter, and selected 1.50 U/μL WarmStart RTx reverse transcriptase (Supplementary Information Figure S4), 0.32 μM Cas12a in combination with 0.16 μM each of Cas12a-guide RNAs (Supplementary Information Figure S5), and 4 µM ssDNA fluorogenic reporter (Supplementary Information Figure S6) for our assay. Upon tuning these reaction conditions, our benchtop DM-compatible CRISPR-Cas12a-assisted RT-RPA could detect 100 µL of 100 GE/µL SARS-CoV-2 RNA, yielding real-time fluorescence amplification curves with onsets of fluorescence increase at ~20 min and strong fluorescence signals after 60 min (Supplementary Information Figure S5 and Figure S6).

### Rapid and sensitive SARS-CoV-2 detection in POC-CRISPR

The development of the benchtop DM-compatible CRISPR-Cas12a-assisted RT-RPA assay allowed us to realize POC-CRISPR with ease, as the assay could be seamlessly adapted in the thermoplastic cartridge and automated by the mobile device. Notably, as the cartridges are fabricated from inexpensive plastic, the material cost for each cartridge is only $0.3 (Supplementary Information Table S2). Upon injecting the mixture of sample and magnetic bead binding buffer in the sample well of the cartridge and mounting the cartridge in the device, the device transported the magnetic beads and bound RNA to the prefilled wash buffer well and then to the prefilled CRISPR-Cas12a-assisted RT-RPA reaction mixture well (**Figure 2A(i)**), and finally commenced reaction incubation and fluorescence measurements. Due to potential differences between the benchtop PCR instrument and the miniature heating module in our mobile DM device, we tuned the reaction temperature and set it at 45.5 °C (Supplementary Information Figure S7). Operating at this reaction temperature, POC-CRISPR could clearly detect 100 µL of 100 GE/µL RNA, as the amplification curves showed onsets of fluorescence increase at ~15 min and steady increases thereafter until 60 min, which were distinct from the slightly decreasing curves in control reactions with no RNA template (**Figure 2A(ii)**). POC-CRISPR could also detect 100 µL of 1 GE/µL RNA, which represents a low viral RNA concentration (**Figure 2A(iii)**). Notably, the amplification curves from 1 GE/µL RNA became distinguishable from the slightly decreasing curves from no RNA controls after only ~30 min, suggesting the possibility of shortening the turnaround time of POC-CRISPR. To our knowledge, these results also represent the first real-time monitoring of a CRISPR-Cas-assisted SARS-CoV-2 detection assay within a POC-amenable device.

**Figure 2.**
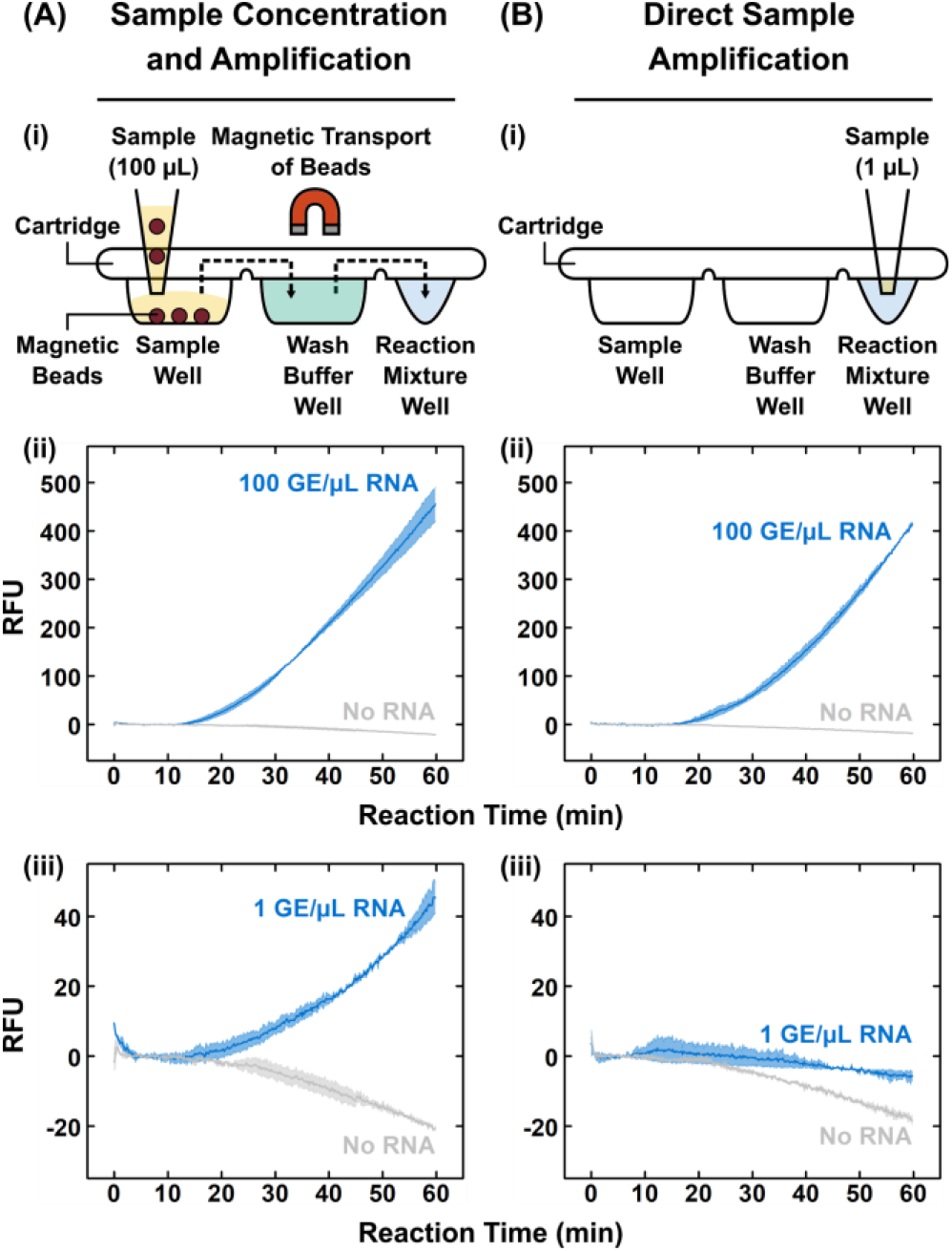
Sensitive detection of SARS-CoV-2 RNA in POC-CRISPR. (A)(i) In POC-CRISPR, magnetic-mediated concentration of RNA from a large sample volume (*e*.*g*., 100 µL) and subsequent automated transfer of magnetic beads and bead-bound RNA to the washer buffer and the reaction mixture within the cartridge enable sensitive detection of SARS-CoV-2. Using POC-CRISPR, (ii) 100 GE/µL RNA samples (blue) can be clearly differentiated from the no RNA controls (gray) and (iii) 1 GE/µL RNA samples (blue) can also be distinguished from the no RNA controls (gray). (B)(i) In contrast, under the more standard “direct sample amplification” scheme, only a small sample volume (*e*.*g*., 1 µL) can be added into the reaction mixture, which can lead to poorer detection sensitivity. (ii) Directly amplified 100 GE/µL RNA samples (blue) can still be clearly differentiated from the no RNA controls (gray), but (iii) directly amplified 1 GE/µL RNA samples (blue) become barely distinguishable from the no RNA controls (gray). Data in (A)(ii), (A)(iii), (B)(ii), and (B)(iii) presented as mean (solid line) ± 1SD (shade), n = 2.

To demonstrate the benefit of sample concentration in POC-CRISPR, we performed comparison experiments in which we directly amplified and detected the sample in the cartridge. Here, we spiked 1 µL SARS-CoV-2 RNA directly into the CRISPR-Cas12a-assisted RT-RPA reaction mixture well of the cartridge (**Figure 2B(i)**). Under direct amplification, samples containing 100 GE/µL RNA were still clearly detected (**Figure 2B(ii)**), though the amplification curves showed slightly slower onsets of fluorescence increase at ~20 min and slightly lower fluorescence signals than those from samples that had been magnetically concentrated. On the other hand, samples containing 1 GE/µL RNA yielded amplification curves that slightly decreased during incubation and appeared similar to control reactions with no RNA template (**Figure 2B(iii)**) noticeably worse than the robust amplification curves from 1 GE/µL RNA samples that had been concentrated. These results illustrate that sample concentration in POC-CRISPR can help detect low concentrations of RNA, which would improve analytical sensitivity and potentially reduce false negatives and enhance clinical sensitivity.

### Evaluation of POC-CRISPR in clinical sample testing

Finally, we evaluated the performance of POC-CRISPR in detecting SARS-CoV-2 directly from clinical NP swab eluates without prior sample processing. We tested 8 samples and compared the results with standard RT-qPCR (with CDC-approved primers and probe^[37]^) performed in a benchtop thermocycler. Here, even for testing these clinical samples, POC-CRISPR allowed us to simply load the mixture of each swab eluate (10 µL) and our magnetic bead buffer into the cartridge, mount the cartridge in the mobile DM device, and initiate testing. POC-CRISPR identified 4 SARS-CoV-2 positive samples that yielded robust fluorescence amplification curves (**Figure 3A**, samples 1 – 4). Notably, the amplification curves for samples 1 and 2 saw onsets of fluorescence increase in 20 min and both saturated the fluorescence detector in the mobile DM device. The 4 samples were also tested as positive by benchtop RT-qPCR, whose cycle of quantification (Cq) values ranged from 22.5 to 26.3. POC-CRISPR also identified 4 SARS-CoV-2 negative samples that yielded slightly decreasing fluorescence signals (**Figure 3A**, samples 5 – 8). These 4 samples were also tested as negative by benchtop RT-qPCR. Moreover, the slightly decreasing fluorescence signals from these 4 negative samples appeared similar to those from the no target control reactions in blank viral transport media (**Figure 3A**, NTC 1 and 2). Overall, POC-CRISPR achieved 8 out of 8 concordance with benchtop RT-qPCR (**Figure 3B**). Importantly, we note that because POC-CRISPR performs real-time fluorescence detection, it has the capacity to report positive results as soon their amplification curves showed onsets of fluorescence increase that allow differentiation from NTCs, thereby shortening the turnaround time. In the case of the 4 positive samples, POC-CRISPR could report these results in 20 – 40 min. Although these 4 positive samples have high viral loads, they nevertheless provide a glimpse for the potential of POC-CRISPR toward rapid SARS-CoV-2 diagnostic testing.

**Figure 3.**
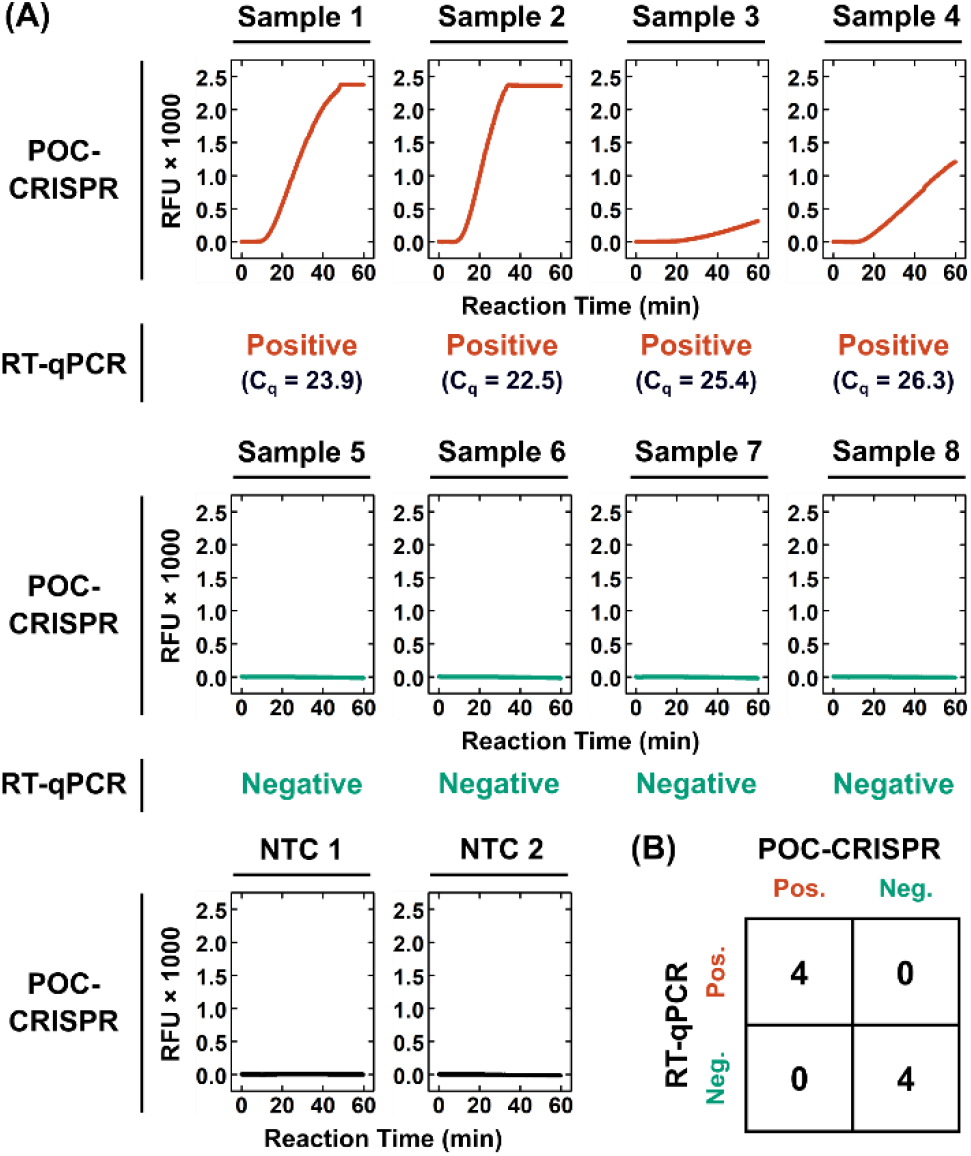
Detection of SARS-CoV-2 from unprocessed clinical nasopharyngeal swab eluates via POC-CRISPR. 8 clinical nasopharyngeal swab eluates are tested by POC-CRISPR without prior sample processing steps, as well as by gold-standard RT-qPCR that uses the CDC-approved primers and probe and is performed in a benchtop thermocycler. (A) POC-CRISPR identifies 4 positive samples that yield robust fluorescence amplification curves (samples 1 – 4), which are tested as positive by benchtop RT-qPCR with cycle of quantification (Cq) values ranging from 22.5 to 26.3. Notably, samples 1 and 2 yield fluorescence signals that saturate the detector in the mobile DM device. POC-CRISPR also identifies 4 negative samples that yield slightly decreasing fluorescence signals (samples 5 – 8) similar to those from the no target control reactions in blank viral transport media (NTC 1 and 2). These 4 samples are also tested as negative by benchtop RT-qPCR. (B) Overall, POC-CRISPR achieves 8 out 8 concordance with benchtop RT-qPCR.

## Conclusions

In summary, we have developed POC-CRISPR, the first CRISPR-Cas-assisted assay that is tenable for POC use. For developing POC-CRISPR, we first developed an integrated benchtop assay by successfully coupling CRISPR-Cas12a-assisted RT-RPA with DM-enabled sample preparation and tuning various assay components (*e*.*g*., reverse transcriptase) and reaction conditions (*e*.*g*., reaction temperature). We subsequently actualized POC-CRISPR by adapting the newly integrated benchtop assay within our custom inexpensive thermoplastic assay cartridge and our palm-sized, integrated mobile DM device, which executed the assay in full automation. We demonstrated that POC-CRISPR is capable of sensitive and rapid detection of SARS-CoV-2 RNA 1 GE/µL from a sample volume of 100 µL in 30 min – which ranks among the most sensitive and fastest CRISPR-Cas-assisted SARS-CoV-2 detection to date. We also evaluated the performance of POC-CRISPR in testing 8 unprocessed clinical NP swab eluates. POC-CRISPR could identify the 4 SARS-CoV-2 positive samples in 20 – 40 min and achieved full concordance with gold-standard benchtop RT-qPCR.

We foresee building upon our promising initial results and taking several routes for improving and expanding POC-CRISPR. First, we foresee improving the analytical sensitivity of POC-CRISPR toward testing clinical samples with low SARS-CoV-2 viral loads. This can be achieved by fine-tuning our magnetic bead buffer so that the input volume of NP swab eluate can be further increased. Second, although POC-CRISPR already involves only a simple manual operation with < 1 min hands-on time, we still envision improving the user-friendliness of POC-CRISPR, which could be accomplished by new cartridge designs that simplify sample injection and reagent prefilling. Third, upon the development of a more sensitive and simpler POC-CRISPR, we can revisit clinical sample testing and focus on samples with low viral loads. Finally, as both DM-enabled sample preparation and CRISPR-Cas-assisted assay can be readily designed for other DNA or RNA targets, we foresee applying POC-CRISPR toward other diseases. Taken together, POC-CRISPR not only represents a significant advance for CRISPR-Cas-assisted diagnostic assays but also has strong potential to become a useful diagnostic tool for COVID-19 and other diseases at the point of care.

## Data Availability

The data that support the findings of this study are available from the corresponding author, KH and THW, upon reasonable request.

## Acknowledgements

The authors are grateful for the financial support from the National Institutes of Health (R01AI138978 and R61AI15462). KH is grateful for the financial support from the Sherrilyn and Ken Fisher Center for Environmental Infectious Diseases at Johns Hopkins University (FCDP-010ZHA2020).

