## Supplementary Information for "Point-of-Care CRISPR-Cas-Assisted SARS-CoV-2 Detection in an Automated and Mobile Droplet Magnetofluidic Device"

### Materials and Methods

Assay reagents

Quantitative Synthetic RNA from SARS-Related Coronavirus 2 (NR-52358) was obtained through the BEI Resources Repository, National Institute of Allergy and Infectious Diseases (NIAID), National Institutes of Health (NIH), and was stored at -80 °C upon receipt. This preparation includes fragments from the open reading frame 1ab (ORF1ab), envelope (E), and nucleocapsid (N) regions.

All oligonucleotides, including RPA primers,[1] Cas12a-guide RNAs,[1] Alexa647-labeled single-stranded DNA (ssDNA) fluorogenic reporter,[2] PCR primers,[3] and fluorescently labeled DNA probe[3] (sequences in Table S1) were purchased from Integrated DNA Technologies (IDT; Coralville, IA). Both Cas12a-guide RNAs were modified with IDT's proprietary 5' AltR1 and 3' AltR2 modifications. Lyophilized Cas12a-guide RNA was reconstituted in DEPC-treated water (Thermo Fisher Scientific, Waltham, MA) at 20 µM. Lyophilized DNA primers, DNA reporter, and DNA probe were reconstituted in nuclease-free water (Promega, Madison, WI) at 100 µM. EnGen Lba (Lachnospiraceae bacterium ND2006) Cas12a (Cpf1) (M0653; 100 µM), WarmStart® RTx reverse transcriptase (M0380; 15000 U/ml), Avian Myeloblastosis Virus (AMV) reverse transcriptase (M0277; 10000 U/mL), and bovine serum albumin (BSA; B9000; 20 mg/mL) were purchased from New England BioLabs (Ipswich, MA). RevertAid reverse transcriptase (EP0441; 200 U/µL) was purchased from Thermo Fisher Scientific (Waltham, MA). TwistAmp Basic kits were purchased from TwistDx Limited (Maidenhead, United Kingdom). qScript XLT 1-Step RT-qPCR ToughMix was purchased from QuantaBio (Beverly, MA). All reconstituted oligonucleotides and aforementioned reagents were stored at -20 °C.

ChargeSwitch™ Total RNA Cell Kit (CS14010) was purchased from Thermo Fisher Scientific (Waltham, MA). Triton™ X-100 was purchased from MilliporeSigma (St. Louis, MO). Both were stored at room temperature.

Assembly of CRISPR-Cas12a-assisted RT-RPA

A typical 20-µL CRISPR-Cas12a-assisted RT-RPA reaction mixture was composed of 1× rehydrated TwistAmp Basic Reaction mix, 0.32 µM each RPA primer, 0.16 µM each Cas12a-guide RNA, 4 µM Alexa647-labeled ssDNA fluorogenic reporter, 0.32 µM EnGen Lba Cas12a, 1.50 U/µL WarmStart RTx reverse transcriptase, 0.01 mg/mL BSA, and 14 mM MgOAc, unless otherwise specified (e.g., in Figures S1 through S6). Inside a PCR hood (AirClean Systems, Creedmoor, NC), assembly began by resuspending each dried pellet of TwistAmp Basic Reaction mix with 29.5 µL rehydration buffer to prepare 1.7× rehydrated TwistAmp Basic Reaction mix. A master mix of all components except WarmStart RTx reverse transcriptase, BSA, and MgOAc were assembled in a 1.5 mL protein low-binding microcentrifuge tube (MilliporeSigma, Burlington, MA) and incubated for 10 min at room temperature to facilitate the formation of Cas12a-guide RNA complexes. WarmStart RTx reverse transcriptase and BSA were then added to the master mix. The master mix was moved from the PCR hood to a biosafety cabinet (The Baker Company, Sanford, ME) to prevent carryover contamination. Inside the biosafety cabinet, the master mix was pipetted into 18-µL MgOAc-free reaction mixture aliquots and then supplemented with 2 µL 140 mM MgOAc to finalize the assembly of the 20-µL CRISPR-Cas12a-assisted RT-RPA reaction mixture.

Benchtop droplet magnetofluidic (DM)-compatible CRISPR-Cas12a-assisted RT-RPA

For preparing the SARS-CoV-2 RNA sample for DM-compatible CRISPR-Cas12a-assisted RT-RPA, SARS-CoV-2 RNA was diluted from the original stock concentration to various experimental titrations by DEPC-treated water. Ten µL diluted SARS-CoV-2 RNA was added to 90 µL 1× PBS to make the 100 µL SARS-CoV-2 RNA sample. The magnetic bead buffer in DM was made by mixing 4 µL of ChargeSwitch magnetic particles (25 mg/ml in 1 mM sodium acetate, pH 4.5) and 10 µL Binding Buffer (B9) from ChargeSwitch™ Total RNA Cell Kit.

DM-compatible CRISPR-Cas12a-assisted RT-RPA began by pipette mixing each 100 µL SARS-CoV-2 RNA sample with 14 µL magnetic bead buffer in a 1.5 mL microcentrifuge tube. The mixture was kept at room temperature for 1 min to allow binding between magnetic beads and SARS-CoV-2 RNA. The tube was placed onto a DynaMag™-2 Magnet (Thermo Fisher Scientific, Waltham, MA) until a visible pellet of magnetic beads was formed on the tube wall before the supernatant was removed via pipetting. Fifty µL Wash Buffer (W14) from ChargeSwitch™ Total RNA Cell Kit was subsequently added into the tube to wash the magnetic beads. The tube was briefly centrifuged and again placed onto the DynaMag™-2 Magnet to pellet the magnetic beads and remove the supernatant. Next, 20 µL CRISPR-Cas12a-assisted RT-RPA reaction mixture was added to the pelleted magnetic beads in each tube. The entire reaction mixture (including the magnetic beads and bound SARS-CoV-2 RNA) was transferred into a PCR tube (0.2 ml 8-Tube PCR Strips, Bio-Rad, Hercules, CA) via pipetting before commencing the reaction in a Bio-Rad CFX96 Touch Real-Time PCR Detection System (Bio-Rad, Hercules, CA) at 43 °C (unless otherwise specified) for 60 min, and the fluorescence signals were measured every 1 min. The fluorescence signals measured by the Bio-Rad CFX96 system were displayed without baseline subtraction (*i.e.*, under "No Baseline Subtraction" mode in the CFX Manager Software). A saturated fluorescence intensity was the maximum intensity which the Real-Time PCR Detection System could determine.

Assay cartridge for POC-CRISPR

The assay cartridge, in which POC-CRISPR was performed, is based on our previous work.[4] Briefly, this cartridge, which houses a sample well, a wash buffer well, and a reaction mixture well, was fabricated from inexpensive plastic components via laser-cutting and thermoforming. The cartridge was composed of 3 layers: a top cap layer that was made of laser-cut polymethylmethacrylate (PMMA) laminated with PTFE tape for establishing a sample injection opening while enclosing the rest of the cartridge, a center spacer layer that was made of laser-cut PMMA laminated with pressure-adhesive tape (PSA, 9472LE adhesive transfer tape, 3M, USA) on both sides for joining the layers, and bottom well layer that was thermoformed from a polypropylene sheet for holding the assay reagents. Upon fabrication of these individual layers, the spacer layer and the well layer were first assembled into an open cartridge. Both the cap layer and the open cartridge were kept at room temperature until use.

Integrated mobile DM device for POC-CRISPR

The integrated mobile DM device, which automated POC-CRISPR in the assay cartridge, is based on from our previous work.[4] Briefly, the device consisted a main housing and a detachable faceplate, both of which were 3D-printed (Formlabs Form 2, black resin) and equipped with permanent neodymium magnets for magnetic clasping and alignment between the two parts. The device housed a motorized magnetic arm, a miniature heating module, and a fluorescence detector (Fluo Sens Integrated, Qiagen), all of which were connected to a microcontroller (Arduino Uno R3) with a motorshield (Arduino Motor Shield Rev3) and an additional custom printed circuit board shield. The motorized magnetic arm located within the main housing of the device. The motorized magnetic arm was 3D-printed, equipped with a pair of permanent neodymium magnets, and actuated by a servo motor (HS-485HB Hitec RCD, Poway, CA, USA) that provided axial movement and a servo motor (PQ12-R Actuonix, Victoria, BC, Canada) that provided linear movement. The movement of the motorized magnetic arm pulled the beads in and out the wells of the assay cartridge, as well as pulled the beads across different wells of the assay cartridge, thereby achieving magnetic transfer within the cartridge. The miniature heating module, located on the detachable faceplate, was an assembly of an aluminum heat block, a thermoelectric cooler, a heat sink and a mini fan. The temperature of the heating module was monitored and controlled by the microcontroller with current supplied by the motorshield.

POC-CRISPR

Prior to performing POC-CRISPR, assay cartridges with pre-loaded reagents were prepared. Specifically, 50 µL wash buffer and 20 µL CRISPR-Cas12a-assisted RT-RPA reaction mixture were loaded into the wash buffer well and the reaction mixture of an open cartridge, respectively. The cap layer was then capped onto the open cartridge before 450 µL silicone oil (50 cSt, Millipore-Sigma, USA) was injected through the sample injection opening to cover both the wash buffer well and the reaction mixture well. The immiscible silicone oil layer above the wash buffer and the reaction mixture served both as a medium for transporting the magnetic beads in the cartridge and as a separator that prevented mixing of assay reagents between the wells. The cartridge was either used immediately or placed on ice with the sample injection opening sealed with tape (Scotch Magic Tape, 3M, USA) until use.

For performing POC-CRISPR, a sample (up to 100 µL in input volume) was pipette mixed with 14 µL magnetic bead buffer and then loaded into the sample well of the assay cartridge. After sample loading, the cartridge was tape sealed and mounted onto the faceplate of the mobile DM device with the reaction mixture well of the assay cartridge seated in the aluminum heat block, whose inner surface was coated with a thermally conductive paste (Arctic Silver 5, Visalia, California, USA) for ensuring consistent thermal contact between the reaction mixture well and the heat block during reaction incubation. The faceplate was then magnetically clipped onto the frame of the device, such that the sample well was positioned between the pair of permanent magnets of the magnetic arm.

POC-CRISPR began with a 2-min automated sample preparation process, during which the magnetic beads and bound RNA were concentrated from the sample well, transferred to the wash buffer well for wash, and finally transferred to the reaction mixture well. Once the magnetic beads and bound RNA arrived in the reaction mixture well, the heating module began heating to initiate CRISPR-Cas-assisted SARS-CoV-2 RT-RPA with real-time fluorescence detection. All reactions were incubated at 45.5 °C (unless otherwise specified) for 60 min. After the heat block reached 45.5 °C, the fluorescence detector (set at a predefined detector current of 65 mA) measured the fluorescence signal from the reaction every 10 s.

Direct SARS-CoV-2 RNA detection in assay cartridge and mobile DM device

For performing direct SARS-CoV-2 detection in the assay cartridge and the mobile DM device, an 18-µL MgOAc-free CRISPR-Cas12a-assisted reaction mixture aliquot supplemented with 1 µL 280 mM MgOAc was added to 1 µL SARS-CoV-2 RNA and then loaded into the reaction mixture of an open cartridge. The cartridge was then capped, injected with silicone oil, and mounted in the mobile DM device. All reactions were then performed under the same conditions as POC-CRISPR.

Clinical sample testing

Eight de-identified clinical nasopharyngeal (NP) swab eluates (i.e., viral transport media) were obtained from the Johns Hopkins Hospital Clinical Microbiology Laboratory in compliance with ethical regulations and the approval of Institutional Review Board (IRB00246027). These clinical NP swab eluates were stored at -80 °C and remained unprocessed (e.g., no RNA extraction nor heat treatment, etc.) until tested by POC-CRISPR. For testing clinical samples, 10 µL unprocessed sample was pipette mixed with a mixture of 14 µL magnetic bead buffer, 90 µL 1× PBS, and 2 µL 10% Triton™ X-100 (for facilitating lysis of viral particles) and loaded into the sample well of an assay cartridge with pre-loaded reagents. Of note, although larger input volumes could be used, only 10 µL of each unprocessed sample was used due to the limited sample volume obtained from the Johns Hopkins Hospital Clinical Microbiology Laboratory. The cartridge was then mounted in the mobile DM device to commence POC-CRISPR. Finally, no RNA template control reactions using 10 µL viral transport medium (R99, Hardy Diagnostics™, Santa Maria, CA) were performed with the same procedure.

In addition to POC-CRISPR, the 8 clinical NP swab eluates were also tested by a RT-qPCR assay with US CDC-approved SARS-CoV-2 N1 primers and probe (Table S1) performed in-house and on benchtop using a Bio-Rad CFX96 Touch Real-Time PCR Detection System. Each 10-µL RT-qPCR reaction was composed of 1× qScript XLT 1-Step RT-qPCR ToughMix, 0.25 µM each primer, 0.25 µM probe, 1 mg/mL BSA, 0.1% Tween-20, and 1 µL of unprocessed clinical sample. RT-qPCR was performed at 50 °C for 10 min (reverse transcription), followed by 95 °C for 30 s and 50 cycles of 95 °C for 5 s and 60 °C for 20 s, with the fluorescence measured at 60 °C of each cycle. The cycle of quantification ($C_q$) of each clinical sample was determined by the Bio-Rad CFX96 system with its built-in regression-based calculation.

Data analysis and presentation

Data were analyzed via Microsoft Excel 365 and plotted in Origin 2018. For benchtop DM-compatible CRISPR-Cas12a-assisted RT-RPA (Figures S1 – S6), the fluorescence signals measured by the Bio-Rad CFX96 system were exported to Excel. For all reactions, the fluorescence signal at 1 min (i.e., $F_{t = 1}$) was subtracted from fluorescence signals at all time points. The subtracted fluorescence signals were subsequently plotted in Origin. For POC-CRISPR using SARS-CoV-2 RNA as the sample (Figures 2 and S7), the fluorescence signals measured by the fluorescence detector in the mobile DM device were first exported to Excel. The fluorescence signals were baseline-corrected by subtracting a linearly fitted line that was calculated from the fluorescence signals acquired between 200 s and 500 s. The mean and standard deviation from 2 technical replicates of baseline-corrected fluorescence signals were calculated in Excel and subsequently plotted in Origin, where the data were presented as mean ± 1SD. For POC-CRISPR clinical sample testing (Figure 3), the fluorescence measurements from the fluorescence detector from each clinical sample were exported and baseline corrected in Excel and subsequently plotted in Origin.

**Table S1. Sequences of RPA primers, Cas12-guide RNAs, fluorogenic reporter, CDC PCR primers, and CDC PCR probe.**

| Component | Sequence (5' → 3') | Reference |
|---|---|---|
| RPA Forward primer | AGGCAGCAGTAGGGGAACTTCTCCTGCTAGAAT | [1] |
| RPA Reverse primer | TTGGCCTTTACCAGACATTTTGCTCTCAAGCTG | |
| Cas12a-guide RNA1 | /AltR1/UAAUUUCUACUAAGUGUAGAU**CAUCACCGCCAUUGCCAGCC**/AltR2/ | |
| Cas12a-guide RNA2 | /AltR1/UAAUUUCUACUAAGUGUAGAU**UUGCUGCUGCUUGACAGAUU**/AltR2/ | |
| Fluorogenic reporter | /Alex647N/TTATT/IAbRQSp/ | [2] |
| CDC PCR N1 Forward primer | GACCCCAAAATCAGCGAAAT | [3] |
| CDC PCR N1 Reverse primer | TCTGGTTACTGCCAGTTGAATCTG | |
| CDC PCR N1 probe | /FAM/ACCCCGCAT/ZEN/TACGTTTGGTGGACC/IABkFQ | |

*Note.* All oligonucleotides synthesized by Integrated DNA Technologies (IDT); Bolded: Target regions in Cas12a-guide RNAs; Abbreviations: Cas12a, New England Biolabs' EnGen Lba Cas12a from *Lachnospiraceae bacterium* ND2006; AltR1 and AltR2, IDT's proprietary Alt-R modifications; IAbRQSp, Iowa Black RQ (quencher); ZEN, internal ZEN quencher; IAbFQSp, Iowa Black FQ (quencher).

**Table S2. Reagents and materials cost breakdown.**

| | Component | Vendor | Amount Per Purchased Unit | | Required Amount Per Cartridge | | Number of Cartridges Per Unit | Unit Price (USD) | Price Per Cartridge (USD) |
|---|---|---|---|---|---|---|---|---|---|
| | | | Value | Unit | Value | Unit | | | |
| **Reagents** | TwistAmp Basic Kit (RPA) | TwistDx | 4.80E+03 | µL | 2.00E+01 | µL | 240 | 410.00 | 1.71 |
| | RPA Forward Primer | IDT | 2.50E+01 | nmol | 6.40E-03 | nmol | 3906 | 15.00 | 0.00 |
| | RPA Reverse Primer | IDT | 2.50E+01 | nmol | 6.40E-03 | nmol | 3906 | 15.00 | 0.00 |
| | Cas12a-guide RNA 1 | IDT | 2.50E+00 | nmol | 3.20E-03 | nmol | 781 | 79.80 | 0.10 |
| | Cas12a-guide RNA 2 | IDT | 2.50E+00 | nmol | 3.20E-03 | nmol | 781 | 79.80 | 0.10 |
| | EnGen Lba Cas12a | NEB | 2.00E+00 | nmol | 6.40E-03 | nmol | 313 | 250.00 | 0.80 |
| | WarmStart RTx RT | NEB | 1.88E+03 | Unit | 3.00E+01 | Unit | 63 | 277.00 | 4.43 |
| | BSA | NEB | 1.20E+01 | mg | 2.00E-04 | mg | 60000 | 32.00 | 0.00 |
| | ChargeSwitch Kit | ThermoFisher Scientific | 2.00E+03 | µL Beads | 4.00E+00 | µL Beads | 500 | 264.00 | 0.53 |
| | Silicone Oil | Millipore Sigma | 2.50E+02 | mL | 4.20E-01 | mL | 595 | 92.50 | 0.16 |
| | | | | | | | | **Reagents Subtotal** | **7.84** |
| **Cartridge Materials** | Top Acrylic Cover | Eplastics | 1.00E+00 | sheet | 1.25E-03 | sheet | 800 | 22.40 | 0.03 |
| | Middle Acrylic Spacer | McMaster | 1.00E+00 | sheet | 8.93E-03 | sheet | 112 | 5.49 | 0.05 |
| | Polypropylene (Thermoformed Wells) | Amazon | 2.00E+01 | folder | 3.13E-02 | folder | 640 | 15.59 | 0.02 |
| | Teflon Tape | McMaster | 5.40E+01 | foot | 5.00E-02 | foot | 1080 | 113.98 | 0.11 |
| | 9472LE 3M Transfer Tape | 3M | 1.92E+02 | foot | 7.14E-02 | foot | 2688 | 280.68 | 0.10 |
| | | | | | | | | **Materials Subtotal** | **0.30** |
| | | | | | | | | **Total** | **8.14** |

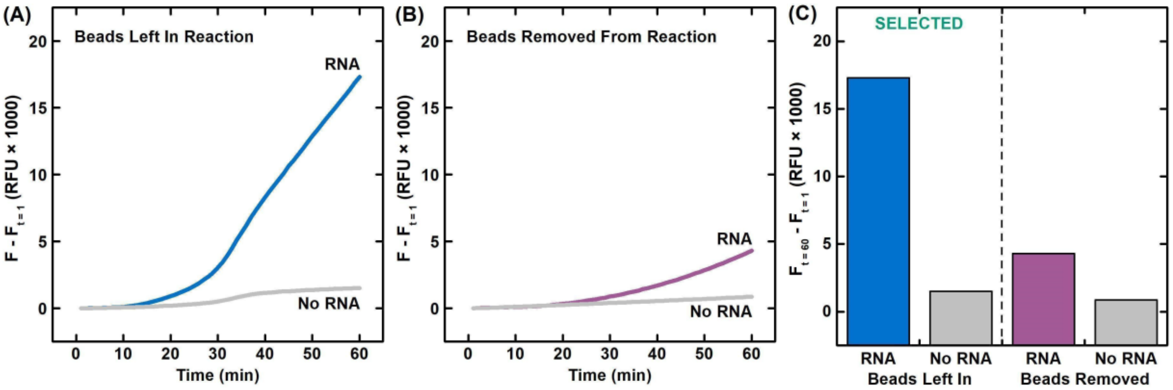


**Figure S1. Initial demonstration of benchtop DM-compatible CRISPR-Cas12a-assisted RT-RPA and comparison of presence of magnetic beads in reaction.** Real-time amplification curves of DM-compatible CRISPR-Cas12a-assisted RT-RPA with (A) the magnetic beads left in the reaction and (B) the magnetic beads removed from the reaction, as well as (C) end point fluorescence signals show that DM-compatible CRISPR-Cas12a-assisted RT-RPA with the magnetic beads left in the reaction can detect SARS-CoV-2 RNA (10000 genome equivalent) with stronger fluorescence signals. Here, the CRISPR-Cas12a-assisted RT-RPA reaction mixture consists of 1× rehydrated TwistAmp Basic Reaction mix, 0.32 µM each of RPA primers, 0.64 µM each of Cas12a-guide RNAs, 4 µM Alexa647-labeled ssDNA fluorogenic reporter, 0.64 µM EnGen Lba Cas12a, 0.75 U/µL WarmStart reverse transcriptase, 0.01 mg/mL BSA, and 14 mM MgOAc. For both conditions, in separate 1.5 mL microcentrifuge tubes, 100 µL synthetic SARS-CoV-2 RNA (100 genome equivalent/µL) and 100 µL nuclease-free water (as the control with no RNA template) are magnetically captured, washed once, and pelleted using a benchtop magnetic rack, before 20 µL CRISPR-Cas12a-assisted RT-RPA reaction mixture is added in each tube and kept at room temperature for ~2 min. For the bead removal condition, the magnetic beads are then pelleted using the benchtop magnetic rack, and the CRISPR-Cas12a-assisted RT-RPA reaction mixture is separated. The 4 CRISPR-Cas12a-assisted RT-RPA reactions are then transferred into separate PCR tubes and performed in a Bio-Rad CFX96 Touch Real-Time PCR Detection System at 43 °C for 60 min, and the fluorescence signals are measured every 1 min. The fluorescence signals measured by the Bio-Rad CFX96 system are displayed without baseline subtraction (*i.e.*, under "No Baseline Subtraction" mode in the CFX Manager Software). Results shown in Figures S2 and S4 – S6 use same benchtop reaction conditions.

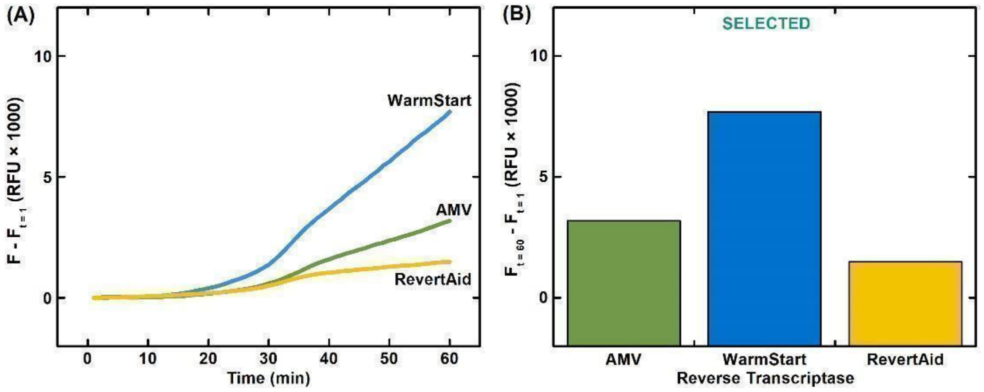


**Figure S2. Comparison of reverse transcriptase in benchtop DM-compatible CRISPR-Cas12a-assisted RT-RPA.** (A) Real-time amplification curves and (B) end-point signals show that, at a fixed dose of 0.75 U/µL, the CRISPR-Cas12a-assisted RT-RPA containing WarmStart RTx reverse transcriptase amplifies magnetically-captured SARS-CoV-2 RNA (10000 genome equivalent) with stronger fluorescence signals than its counterparts containing either AMV (Avian Myeloblastosis Virus) reverse transcriptase or RevertAid reverse transcriptase. Here, the CRISPR-Cas12a-assisted RT-RPA reaction mixture consists of 1× rehydrated TwistAmp Basic Reaction mix, 0.32 µM each of RPA primers, 0.64 µM each of Cas12a-guide RNAs, 4 µM Alexa647-labeled ssDNA fluorogenic reporter, 0.64 µM EnGen Lba Cas12a, 0.75 U/µL of either WarmStart, AMV, or RevertAid reverse transcriptase, 0.01 mg/mL BSA, and 14 mM MgOAc. For each of the 3 reaction conditions, in separate 1.5 mL microcentrifuge tubes, 100 µL synthetic SARS-CoV-2 RNA (100 genome equivalent/µL) is magnetically captured, washed once, and pelleted using a benchtop magnetic rack, before 20 µL CRISPR-Cas12a-assisted RT-RPA reaction mixture is added in each tube. The 3 CRISPR-Cas12a-assisted RT-RPA reactions are then transferred into separate PCR tubes and performed in a Bio-Rad CFX96 Touch Real-Time PCR Detection System with the conditions described in Figure S1.

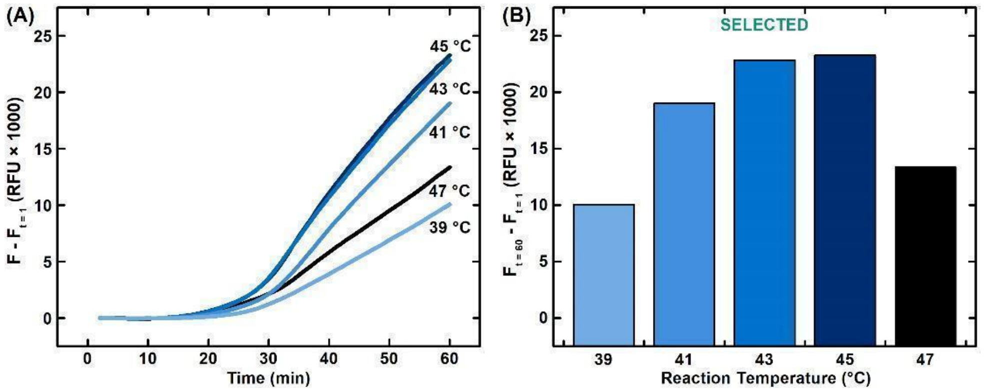


**Figure S3. Comparison of reaction temperature for benchtop DM-compatible CRISPR-Cas12a-assisted RT-RPA.** (A) Real-time amplification curves and (B) end-point signals show that, when performed at either 43 °C or 45 °C, CRISPR-Cas12a-assisted RT-RPA amplifies magnetically-captured SARS-CoV-2 RNA (10000 genome equivalent) with stronger fluorescence signals than their counterparts performed 39 °C, 41 °C, or 47 °C. Here, the CRISPR-Cas12a-assisted RT-RPA reaction mixture consists of 1× rehydrated TwistAmp Basic Reaction mix, 0.32 µM each of RPA primers, 0.64 µM each of Cas12a-guide RNAs, 4 µM Alexa647-labeled ssDNA fluorogenic reporter, 0.64 µM EnGen Lba Cas12a, 0.75 U/µL WarmStart RTx reverse transcriptase, 0.01 mg/mL BSA, and 14 mM MgOAc. For each of the 5 reaction conditions, 100 µL synthetic SARS-CoV-2 RNA (100 genome equivalent/µL) is magnetically captured, washed once, and pelleted using a benchtop magnetic rack, before 20 µL CRISPR-Cas12a-assisted RT-RPA reaction mixture is added in each tube. The 5 CRISPR-Cas12a-assisted RT-RPA reactions are then transferred into separate PCR tubes and performed in a Bio-Rad CFX96 Touch Real-Time PCR Detection System at the specified temperatures (using the built-in temperature gradient function) for 60 min, and the fluorescence signals are measured every 1 min. The fluorescence signals measured by the Bio-Rad CFX96 system are displayed without baseline subtraction (*i.e.*, under "No Baseline Subtraction" mode in the CFX Manager Software).

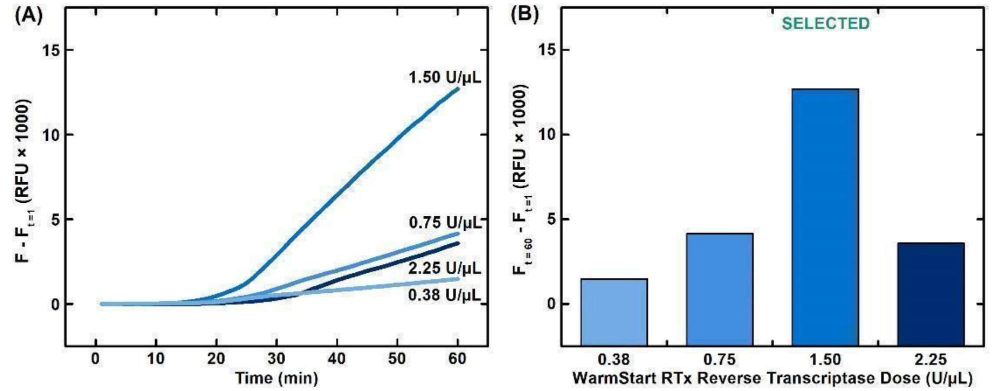


**Figure S4. Comparison of WarmStart RTx reverse transcriptase dose in benchtop DM-compatible CRISPR-Cas12a-assisted RT-RPA.** (A) Real-time amplification curves and (B) end-point signals show that, the CRISPR-Cas12a-assisted RT-RPA reaction containing 1.50 U/µL WarmStart RTx reverse transcriptase amplifies magnetically-captured SARS-CoV-2 RNA (10000 genome equivalent) with stronger fluorescence signals than its counterparts containing 0.38 U/µL, 0.75 U/µL, and 2.25 U/µL. Here, the CRISPR-Cas12a-assisted RT-RPA reaction mixture consists of 1× rehydrated TwistAmp Basic Reaction mix, 0.32 µM each of RPA primers, 0.64 µM each of Cas12a-guide RNAs, 4 µM Alexa647-labeled ssDNA fluorogenic reporter, 0.64 µM EnGen Lba Cas12a, WarmStart RTx reverse transcriptase at the specified doses, 0.01 mg/mL BSA, and 14 mM MgOAc. For each of the 4 reaction conditions, in separate 1.5 mL microcentrifuge tubes, 100 µL synthetic SARS-CoV-2 RNA (100 genome equivalent/µL) is magnetically captured, washed once, and pelleted using a benchtop magnetic rack, before 20 µL CRISPR-Cas12a-assisted RT-RPA reaction mixture is added in each tube. The 4 CRISPR-Cas12a-assisted RT-RPA reactions are then transferred into separate PCR tubes and performed in a Bio-Rad CFX96 Touch Real-Time PCR Detection System with the conditions described in Figure S1.

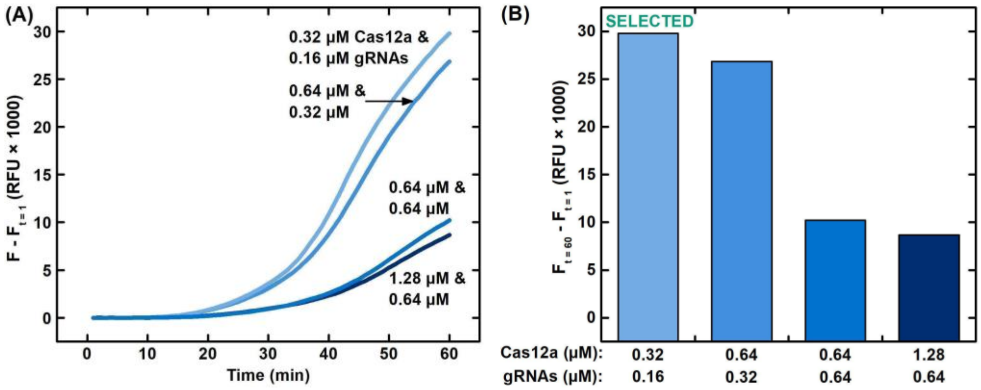


**Figure S5. Comparison of Cas12a and Cas12a-guide RNA concentrations in benchtop DM-compatible CRISPR-Cas12a-assisted RT-RPA.** (A) Real-time amplification curves and (B) end-point signals show that, the CRISPR-Cas12a-assisted RT-RPA reaction containing 0.32 µM EnGen Lba Cas12a and 0.16 µM each of Cas12a-guide RNAs amplifies magnetically-captured SARS-CoV-2 RNA (10000 genome equivalent) with stronger fluorescence signals than its counterparts containing 0.64 µM Cas12a and 0.32 µM guide RNAs, 0.64 µM Cas12a and 0.64 µM guide RNAs, and 1.28 µM Cas12a and 0.64 µM guide RNAs. Here, the CRISPR-Cas12a-assisted RT-RPA reaction mixture consists of 1× rehydrated TwistAmp Basic Reaction mix, 0.32 µM each of RPA primers, each of Cas12a-guide RNAs at the specified concentrations, 4 µM Alexa647-labeled ssDNA fluorogenic reporter, EnGen Lba Cas12a at their specified concentrations, 1.50 U/µL WarmStart RTx reverse transcriptase, 0.01 mg/mL BSA, and 14 mM MgOAc. For each of the 4 reaction conditions, in separate 1.5 mL microcentrifuge tubes, 100 µL synthetic SARS-CoV-2 RNA (100 genome equivalent/µL) is magnetically captured, washed once, and pelleted using a benchtop magnetic rack, before 20 µL CRISPR-Cas12a-assisted RT-RPA reaction mixture is added in each tube. The 4 CRISPR-Cas12a-assisted RT-RPA reactions are then transferred into separate PCR tubes and performed in a Bio-Rad CFX96 Touch Real-Time PCR Detection System with the conditions described in Figure S1.

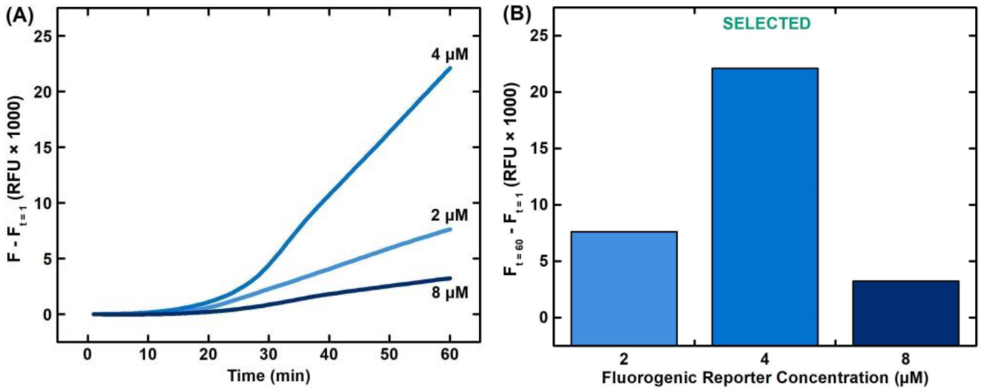


**Figure S6. Comparison of fluorogenic reporter concentration in benchtop DM-compatible CRISPR-Cas12a-assisted RT-RPA.** (A) Real-time amplification curves and (B) end-point signals show that, the CRISPR-Cas-assisted RT-RPA reaction containing 4 µM Alexa647-labeled ssDNA fluorogenic reporter amplifies magnetically-captured SARS-CoV-2 RNA (10000 genome equivalent) with stronger fluorescence signals than its counterparts containing 2 µM fluorogenic reporter and 8 µM fluorogenic reporter. Here, the CRISPR-Cas12a-assisted RT-RPA reaction mixture consists of 1× rehydrated TwistAmp Basic Reaction mix, 0.32 µM each of RPA primers, 0.16 µM each of Cas12a-guide RNAs, Alexa647-labeled ssDNA fluorogenic reporter at the specified concentrations, 0.32 µM EnGen Lba Cas12a, 1.50 U/µL WarmStart RTx reverse transcriptase, 0.01 mg/mL BSA, and 14 mM MgOAc. For each of the 3 reaction conditions, in separate 1.5 mL microcentrifuge tubes, 100 µL synthetic SARS-CoV-2 RNA (100 genome equivalent/µL) is magnetically captured, washed once, and pelleted using a benchtop magnetic rack, before 20 µL CRISPR-Cas-assisted RT-RPA reaction mixture is added in each tube. The 3 CRISPR-Cas12a-assisted RT-RPA reactions are then transferred into separate PCR tubes and performed in a Bio-Rad CFX96 Touch Real-Time PCR Detection System with the conditions described in Figure S1.

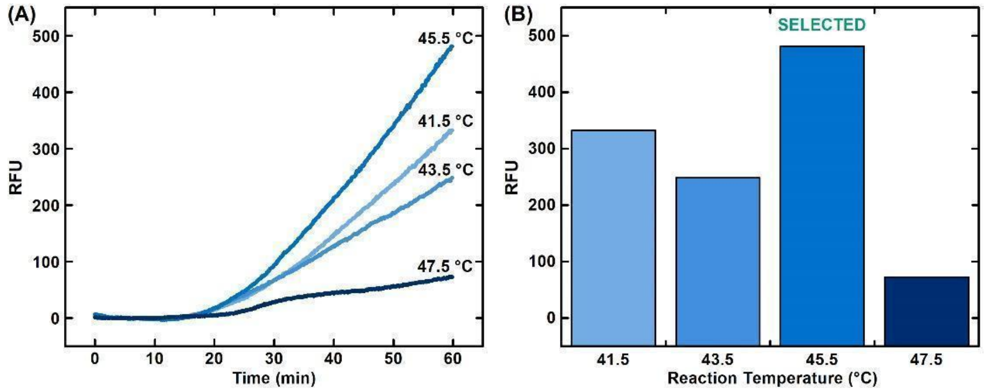


**Figure S7. Comparison of reaction temperature for POC-CRISPR.** POC-CRISPR is realized by adapting the benchtop DM-compatible CRISPR-Cas12a-assisted RT-RPA assay within a custom thermoplastic assay cartridge and an integrated mobile DM device that executes the assay steps in the cartridge in full automation. (A) Real-time amplification curves and (B) end-point signals show that, when performed at 45.5 °C, POC-CRISPR detects SARS-CoV-2 RNA (10000 genome equivalent) with stronger fluorescence signals than its counterparts performed 41.5 °C, 43.5 °C, or 47.5 °C. Here, the CRISPR-Cas12a-assisted RT-RPA reaction mixture consists of 1× rehydrated TwistAmp Basic Reaction mix, 0.32 µM each of RPA primers, 0.16 µM each of Cas12a-guide RNAs, 4 µM Alexa647-labeled ssDNA fluorogenic reporter, 0.32 µM EnGen Lba Cas12a, 1.50 U/µL WarmStart RTx reverse transcriptase, 0.01 mg/mL BSA, and 14 mM MgOAc. For each of the 4 reaction conditions, in separate cartridges, 100 µL synthetic SARS-CoV-2 RNA (100 genome equivalent/µL) is loaded. Each cartridge is mounted in the mobile DM device to magnetically capture RNA, wash, and transfer the RNA into the CRISPR-Cas12a-assisted RT-RPA reaction mixture. The miniature heater in the device then heats the reaction well in each cartridge to the specified temperature for 60 min while the fluorescence detector in the device measures the fluorescence signal every 10 s. The fluorescence signals measured by the fluorescence detector are custom baseline corrected, where a linearly fitted line from fluorescence signals between 200 s and 500 s is subtracted from the measured fluorescence signal at each time point.
